# Clinical outcomes of chimeric antigen receptor T cell therapy in 21 patients with Relapse/Refractory Ileocecal Region B cell lymphoma

**DOI:** 10.1101/2025.08.26.25334457

**Authors:** Zeyan Shi, Xing Chen, Fankai Meng, Yang Cao, Zhicheng Zhang, Zhenfang Liu, Wang Wei, Xiaojian Zhu

**Affiliations:** Department of Hematology, the First Affiliated Hospital of Guangxi Medical University, Nanning, China; Department of Hematology, Tongji Hospital, Tongji Medical College, Huazhong University of Science and Technology, Wuhan 430030, Hubei, China; Department of Gastroenterology, Tongji Hospital, Tongji Medical College, Huazhong University of Science and Technology, Wuhan 430030, Hubei, China; Immunotherapy Research Center for Hematologic Diseases of Hubei Province, Wuhan 430030, Hubei, China

**Keywords:** Ileocecal Valve, CAR-T cell therapy, Lymphoma B-Cell Progression-Free Survival, Recurrence

## Abstract

**Objective:** To evaluate the efficacy, safety, and prognostic factors of chimeric antigen receptor T-cell (CAR-T) therapy in patients with relapsed/refractory ileocecal region lymphoma and provide evidence for salvage therapy.

**Methods:** A retrospective analysis was conducted on 21 ileocecal region lymphoma patients (IC group) and matched 23 non-ileocecal lymphoma patients (non-IC group) treated with CAR-T therapy between June 2014 and August 2024. Baseline characteristics, genetic mutations (next-generation sequencing), CAR-T cell kinetics (digital PCR), treatment response (WHO criteria), overall survival (OS), and progression-free survival (PFS) were assessed. Kaplan-Meier analysis, Cox regression, and Lasso models were used to identify prognostic factors.

**Results:** The IC group showed significantly lower 3-month objective response rate (ORR) (55.56% versus 86.96%, *P=*0.036) and shorter median PFS (3.5 months versus 27 months, *P=*0.0003) compared to the non-IC group, but no OS difference (*P=*0.766). CAR-T cell expansion was higher in the IC group (CD19 C_max_: 67,290 versus 35,400, *P=*0.009), but persistence was similar. TP53 mutations (42.86% in IC group) showed no significant correlation with treatment response. Safety profiles (cytokine release syndrome [CRS] and immune effector cell-associated neurotoxicity syndrome[ICANS]) were comparable between groups (*P*>0.05). Multivariate analysis identified poor 3-month treatment response (HR=32.075, *P=*0.015) and higher pre-treatment lines (HR=8.6, *P=*0.036) as independent risk factors for PFS. Delayed B-cell recovery and low baseline lymphocyte counts were associated with worse OS.

**Conclusion:** CAR-T therapy is feasible and safe for relapsed/refractory ileocecal lymphoma, but short-term efficacy (ORR, PFS) is inferior to non-ileocecal patients. High CAR-T expansion did not improve long-term outcomes. Early response assessment (3 months), baseline lymphocyte levels, and B-cell recovery are critical prognostic indicators, suggesting the need for optimized timing and combination strategies to enhance efficacy.

## INTRODUCTION

The ileocecal region is the most commonly involved site in primary intestinal lymphoma, accounting for approximately 20.3-39.8% of all cases ^[1-4]^. The predominant pathological types include diffuse large B-cell lymphoma (DLBCL) and mucosa-associated lymphoid tissue (MALT) lymphoma^[5]^. Ileocecal lymphoma frequently presents with acute complications such as abdominal pain, intestinal obstruction, perforation, or hemorrhage^[3]^, which may necessitate emergency surgical intervention and disrupt the continuity of systemic antitumor therapy. Currently, first-line treatment for ileocecal lymphoma primarily relies on R-CHOP chemotherapy; however, therapeutic efficacy varies significantly due to biological heterogeneity, with approximately 20-30% of patients experiencing disease relapse or progression^[2]^. For these patients, traditional salvage therapies, such as autologous hematopoietic stem cell transplantation (ASCT) combined with high-dose chemotherapy, yield limited benefits.In recent years, chimeric antigen receptor T-cell (CAR-T) therapy has emerged as a revolutionary approach for relapsed/refractory B-cell lymphomas. Early-phase ZUMA-1 studies demonstrated a 58% complete remission rate with CAR-T therapy, significantly surpassing outcomes from salvage ASCT alone^[6]^. Furthermore, CAR-T therapy has shown favorable efficacy and safety in relapsed/refractory primary gastrointestinal lymphomas^[7]^. Nevertheless, no dedicated studies have specifically addressed CAR-T therapy for ileocecal lymphoma. This study focuses on 21 patients with ileocecal lymphoma treated with CAR-T therapy. By integrating molecular profiling and systematically analyzing associations with overall survival (OS), progression-free survival (PFS), and treatment-related toxicities, we aim to establish a theoretical framework for personalized CAR-T therapy in this distinct patient subgroup.

## METHODS

### Patients

We retrospectively analyzed 21 patients with histologically or radiologically confirmed ileocecal lymphoma treated at Tongji Hospital, Tongji Medical College, Huazhong University of Science and Technology between June 2014 and August 2024. A propensity score-matched cohort of 23 non-ileocecal lymphoma patients served as controls. Ileocecal involvement was defined as disease affecting the ileocecal valve, terminal ileum, cecum, or appendix. The patients were staged according to Lugano staging system for gastrointestinal lymphoma, and the Ann Arbor staging system was used for non-ileocecal patients. Each patient completed the following staging examinations: physical examination, CT plain scan enhancement or PET/CT, bone marrow cytology, and bone marrow biopsy. B symptoms were defined as the presence of at least one of the following: unexplained recurrent fever (temperature ≥38°C), night sweats, or weight loss (≥10% body weight within 6 months).

### Data Collection

Baseline assessments included demographic characteristics, serological/immunological parameters, treatment-related adverse events, and radiographic findings. Treatment response was evaluated at 3 months post-CAR-T using contrast-enhanced CT or PET/CT.

### Histology

Formalin-fixed biopsy specimens were histologically classified according to WHO Classification of Haematolymphoid Tumours (5th edition). Immuno-histochemical staining included LCA, CD20, CD19, CD22, CD3, CD5, CD10, CD30, CD45RO, CD56, ALK, BCL-2, BCL-6, C-myc, and Cyclin D1. Double-expressor lymphoma was defined as co-expression of C-myc with either BCL-2 or BCL-6 by immuno-histochemistry.

### Molecular Genetics

Double-expressor cases underwent fluorescence in situ hybridization (FISH) to confirm double-hit lymphoma (C-myc with BCL-2 and/or BCL-6 rearrangements). Gene mutation profiling was performed using next-generation sequencing (NGS) or PCR-based methods.

### Therapy Procedure

Among the 21 patients, 11 received autologous stem cell transplantation (ASCT) followed by CD19/20/22 CAR-T cell therapy, with prior collection of both autologous stem cells and peripheral blood mononuclear cells (PBMCs). The remaining 10 patients underwent CD19/22 CAR-T cell therapy alone following PBMC collection. CAR-T cell products were manufactured. Bridging therapy was administered when clinically indicated during CAR-T cell manufacturing, with regimen selection guided by institutional experience, disease status, and prior treatment history. Conditioning regimens for the ASCT-CAR-T cohort included BEAM (carmustine, etoposide, cytarabine, melphalan), BEAMF (BEAM plus fludarabine), or liposomal doxorubicin combined with BEAM, while the CAR-T-only cohort received fludarabine/cyclophosphamide (FC). Patients subsequently received infusions of commercially available CD19/CD22 CAR-T products (Relma-cel or Axi-cel) or investigational CD19/CD22 CAR-T cells within 2-6 days post-conditioning, either as single or sequential infusions.

Among 23 non-ileocecal patients, 17 received ASCT followed by CAR-T cell therapy. The ASCT conditioning regimens included BEAM (9 cases), doxorubicin + BEAM (4 cases), BEAMF (1 case), and TBC (1 case). Patients in the CAR-T group received the FC regimen as conditioning chemotherapy, with the infusion procedure consistent with that of the ileocecal group.

Both ileocecal and non-ileocecal patients were administered structurally identical CAR-T cells provided by Senlang Bio.

### Response Criteria

Treatment response was assessed using WHO criteria (complete response [CR], partial response [PR], stable disease [SD], progressive disease [PD]). Initial response evaluation occurred at 3 months post-infusion. Overall survival (OS) was defined from CAR-T infusion to death from any cause or last follow-up (March 7, 2025). Progression-free survival (PFS) spanned from infusion to relapse, progression, death, or last follow-up. CRS and ICANS were graded per ASTCT consensus criteria ^[8]^.

### Statistical Analysis

Categorical variables were compared using Fisher’s exact test; continuous variables with t-test. Survival curves were generated via Kaplan-Meier method with log-rank testing. Lasso regression model was used for multivariate analysis of PFS, and Logistic regression model was used for multivariate analysis of OS. The significant and clinically significant variables in univariate analysis were included. GraphPad Prism 10.0 analyzed clinical characteristics and survival curves; SPSS 29.0 (SPSS Inc, Chicago, IL) performed multivariate analyses.

## RESULTS

### Patient Characteristics

The clinical characteristics of the 21 patients are summarized in **Table 1**. Prior to CAR-T therapy, patients had received a mean of 2.381 lines of treatment (range: 1-5). The median age was 45 years (range: 21-71), with a male predominance (80.95%, 17/21; male-to-female ratio: 4.25:1). All cases were of B-cell origin, with diffuse large B-cell lymphoma (DLBCL) being the most common histology (85.71%, 18/21). Among DLBCL cases, 10 (55.56%) were non-germinal center B-cell-like (non-GCB) subtype and 8 (44.44%) were germinal center B-cell-like (GCB) subtype.

**Table 1.**
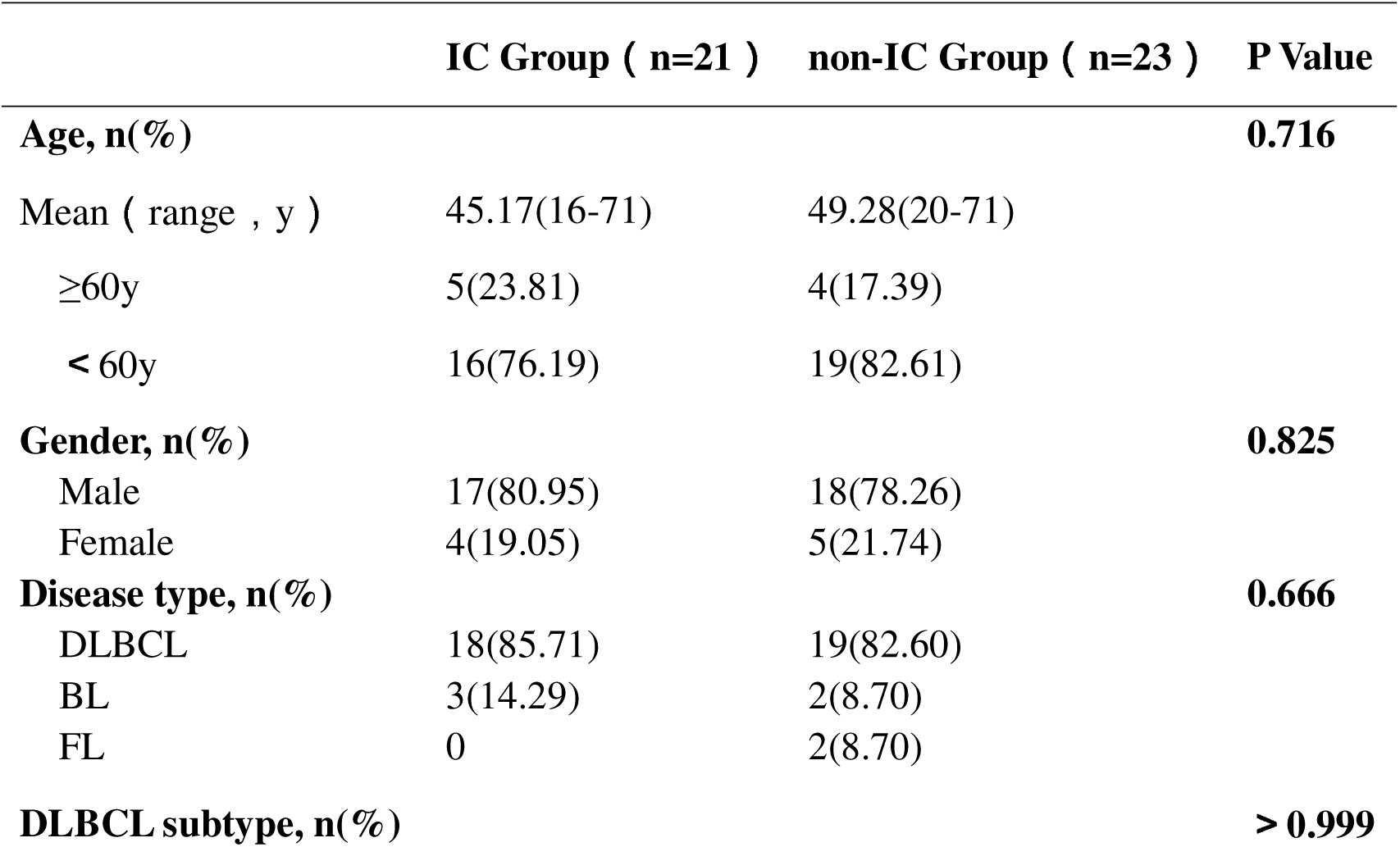
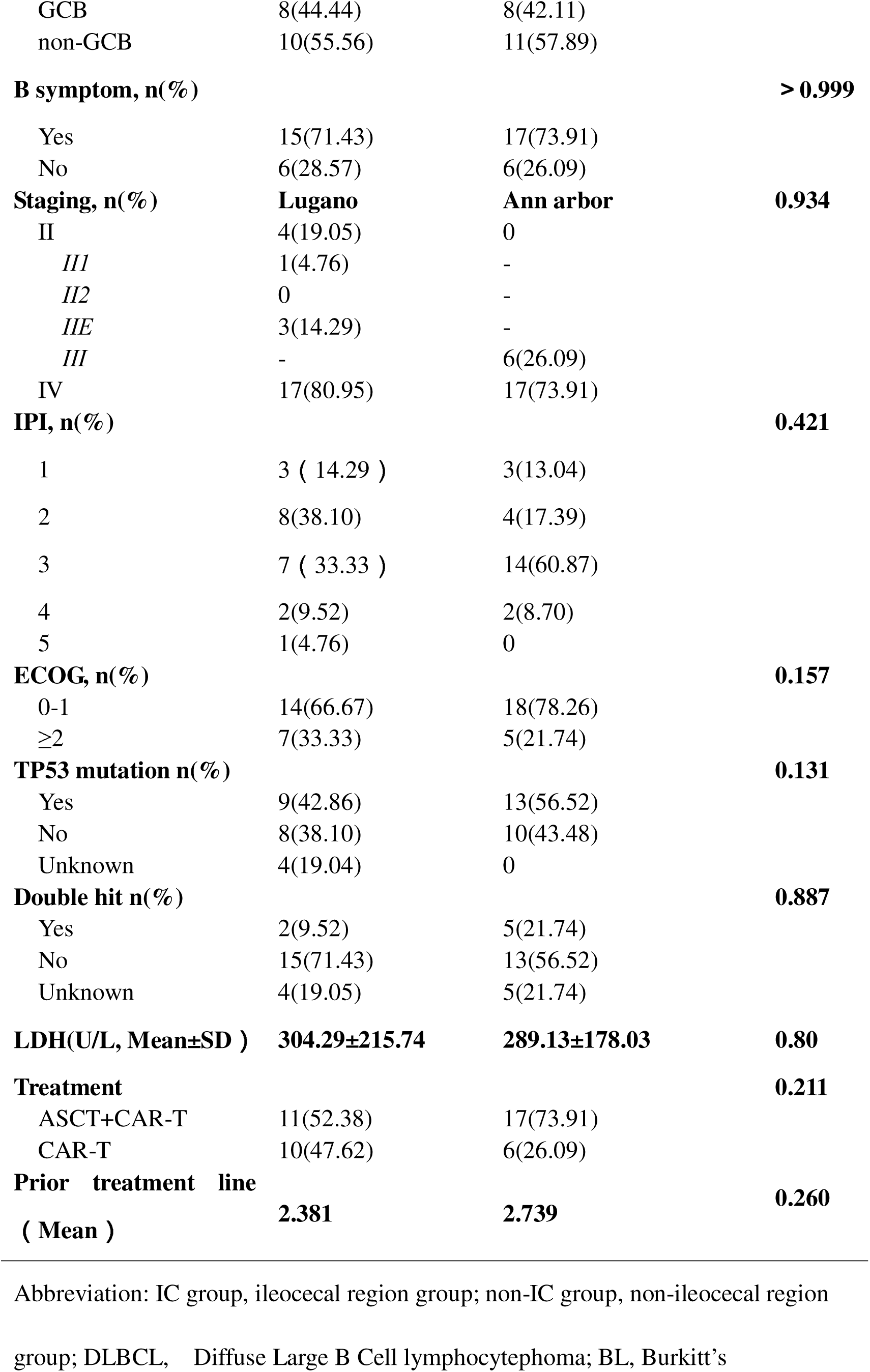
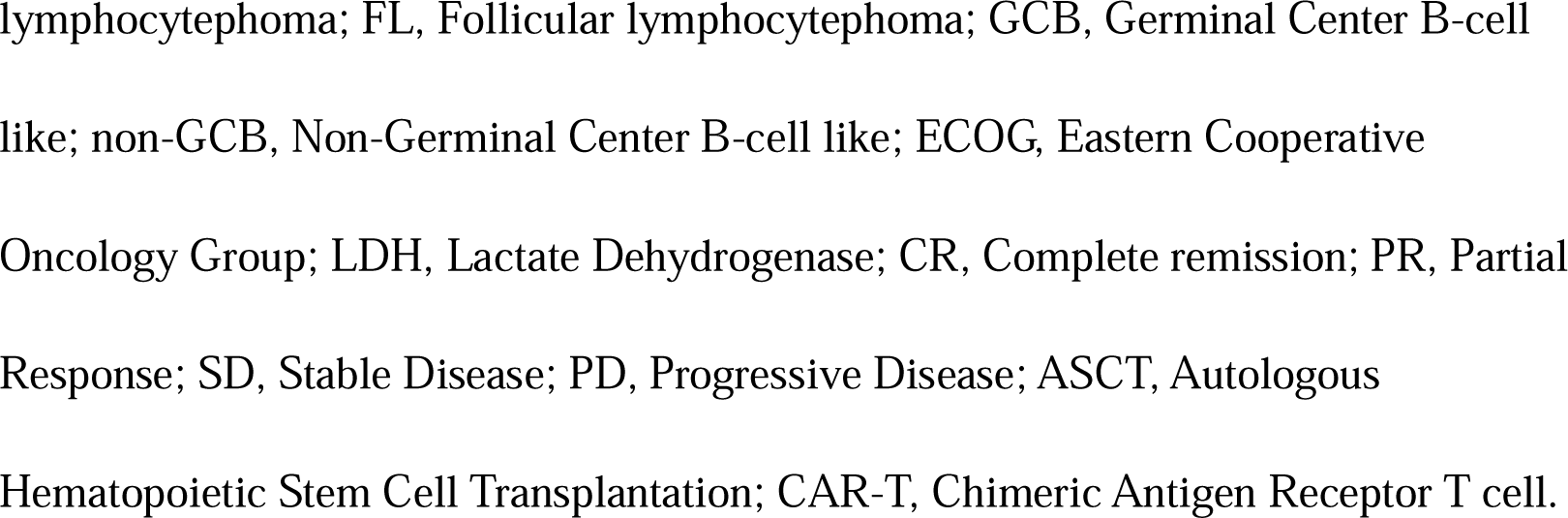
Baseline characteristics of ileocecal (IC) and non-ileocecal (non-IC) lymphoma patients.

At initial diagnosis, 15 patients (71.43%) presented with B symptoms. According to Lugano staging, 17 patients (80.95%) had stage IV disease at onset, including 6 cases with distant extranodal involvement. The most common non-gastrointestinal sites were bone marrow (3 cases) and bone (2 cases), followed by central nervous system, vertebrae, and pleura (1 case each). Three patients (14.29%) had stage IIE disease, and 1 (4.76%) had stage II1 disease. Most patients had intermediate-risk International Prognostic Index (IPI) scores (2-3 points)Molecular Genetic Characteristics was shown in **Fig. 1**. Fifteen patients underwent genetic mutation profiling, revealing a mean of 4.8 mutations per patient (range: 1-12). TP53 mutations were the most frequent alteration, detected in 9 ileocecal lymphoma patients (60%). These frequently co-occurred with CD79B, KMT2D, TET2, and EP300 mutations. BTG2 mutations were present in 3 patients (20%).

**Fig. 1.**
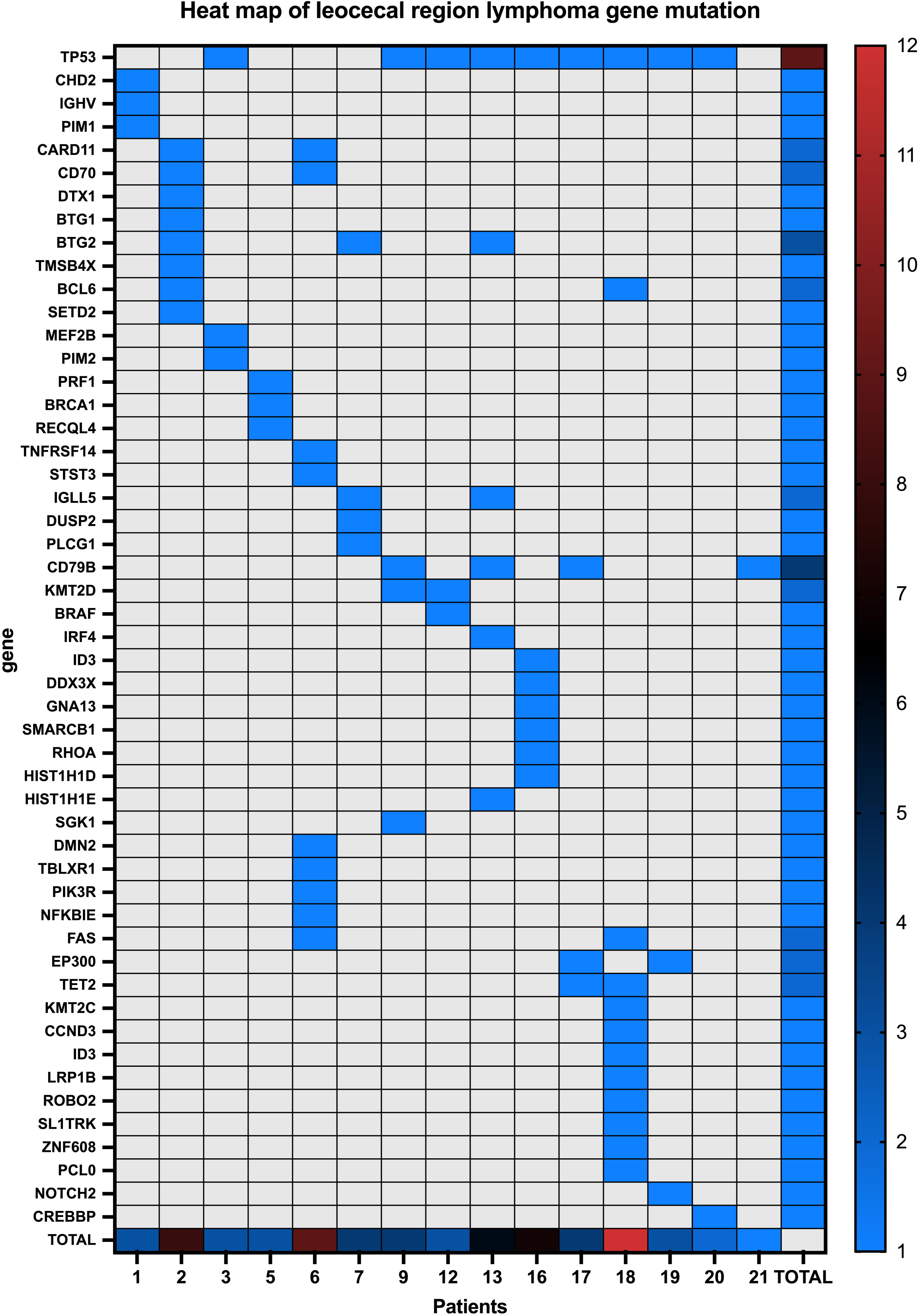
Mutation profile in ileocecal lymphoma. This heatmap illustrates the distribution of high-frequency mutations among ileocecal lymphoma patients. The x-axis represents patient identifiers (n=15), while the y-axis lists candidate genes. The blue gradient indicates mutation frequencies across patients, with darker shades representing higher frequencies. Right-side numerals (1-12) denote mutation frequency rankings (ascending order). The "TOTAL" row and column display aggregate mutation frequencies across the cohort.

At initial presentation, 14 patients (66.67%) reported gastrointestinal symptoms as their chief complaint **(Fig.2-A)**. The most common manifestations were abdominal pain and intestinal obstruction (5 cases each), followed by abdominal distension and altered bowel habits (4 cases each). Four patients required surgical intervention for intestinal complications.

**Fig. 2.**
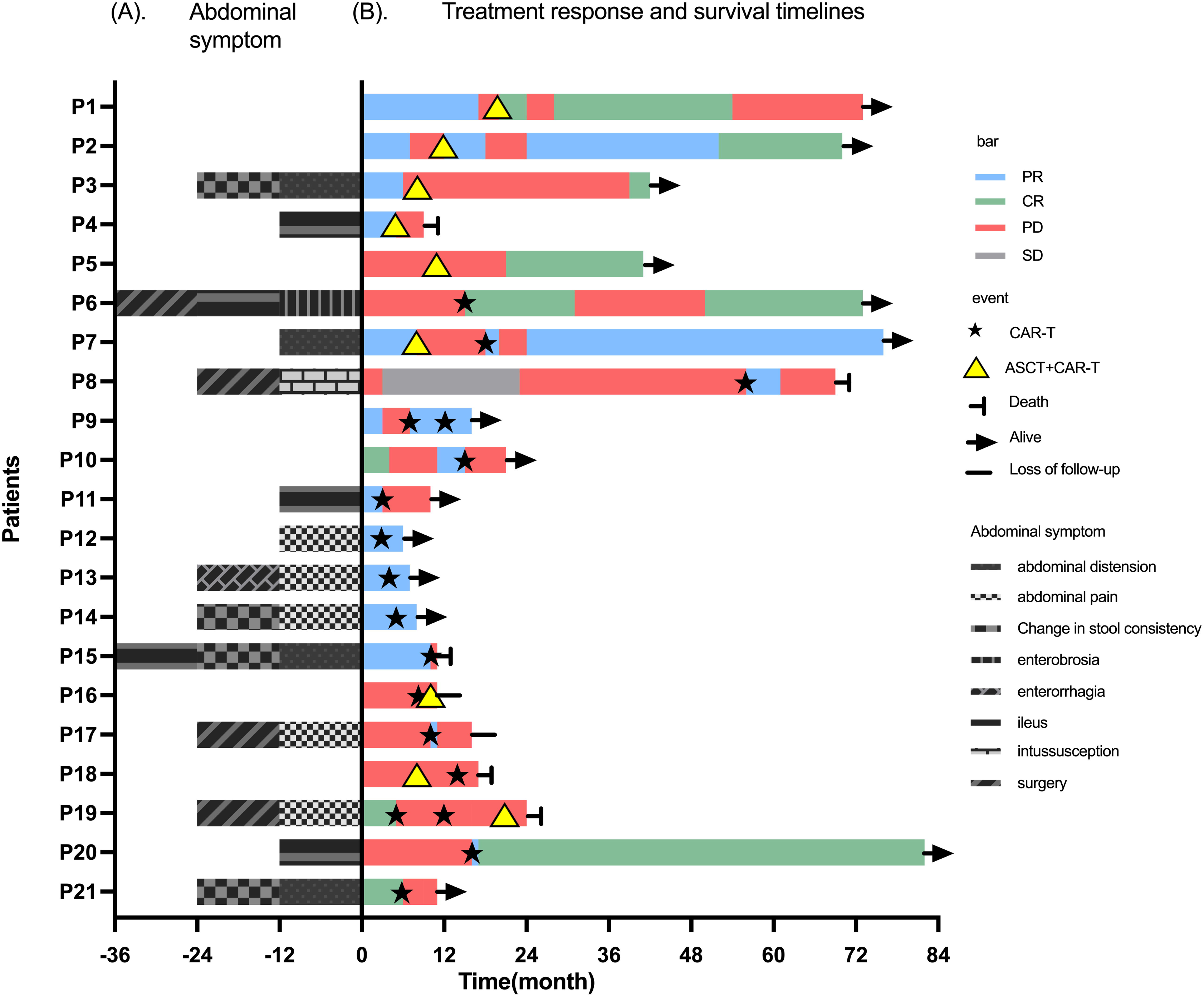
Abdominal symptoms and treatment responses. **(A)** Abdominal symptoms at diagnosis. This figure illustrates the clinical distribution of abdominal symptoms in ileocecal lymphoma patients. The x-axis represents symptom categories, and the y-axis displays the number of symptomatic patients (range: 0-6). (**B)** Treatment response and survival timeline. This swimmer plot depicts the temporal evolution of disease status and treatment responses. The x-axis represents follow-up duration (0-84 months), and the y-axis lists patient identifiers (P1-P21). Horizontal lines denote individual patient trajectories, with colors/symbols marking key clinical events. Five patients (P8, P19, P18, P15, P4) died before study termination (data censored), and one patient (P17) was lost to follow-up. CR: Complete remission; PR: Partial Response; SD: Stable Disease; PD: Progressive Disease; ASCT: Autologous Hematopoietic Stem Cell Transplantation; CAR-T: Chimeric Antigen Receptor T cell therapy.

### Objective Response

Among the 21 patients **(Fig.2-B)**, 11 received autologous hematopoietic stem cell transplantation (ASCT) followed by CAR-T therapy, while 10 underwent CAR-T therapy alone. By March 7, 2025, 18 patients (excluding 3 with <3 months of follow-up) completed 3-month response assessments**(Fig.3)**. In the ileocecal subgroup, 3 patients achieved complete remission (CR), 6 achieved partial remission (PR), resulting in an objective response rate (ORR) of 55.56% (10/18; 95% CI: 38.11-94.35), which was significantly lower than the non-ileocecal group (86.96%, *P=*0.036). Among responders (n=9), the maximum response duration was 62 months; however, 7 patients (3 CR, 4 PR) experienced relapse or progression at a median of 7 months (range:3-21). One patient achieved sustained partial remission after secondary CD19/CD22 CAR-T retreatment. Subgroup analyses revealed no significant differences in ORR based on abdominal symptom presence (63.64% versus 42.86%, *P=*0.631), TP53 mutation status (57.14% versus 71.43%, P > 0.999), or ASCT bridging (44.44% versus 66.67%, *P=*0.637).

**Fig. 3.**
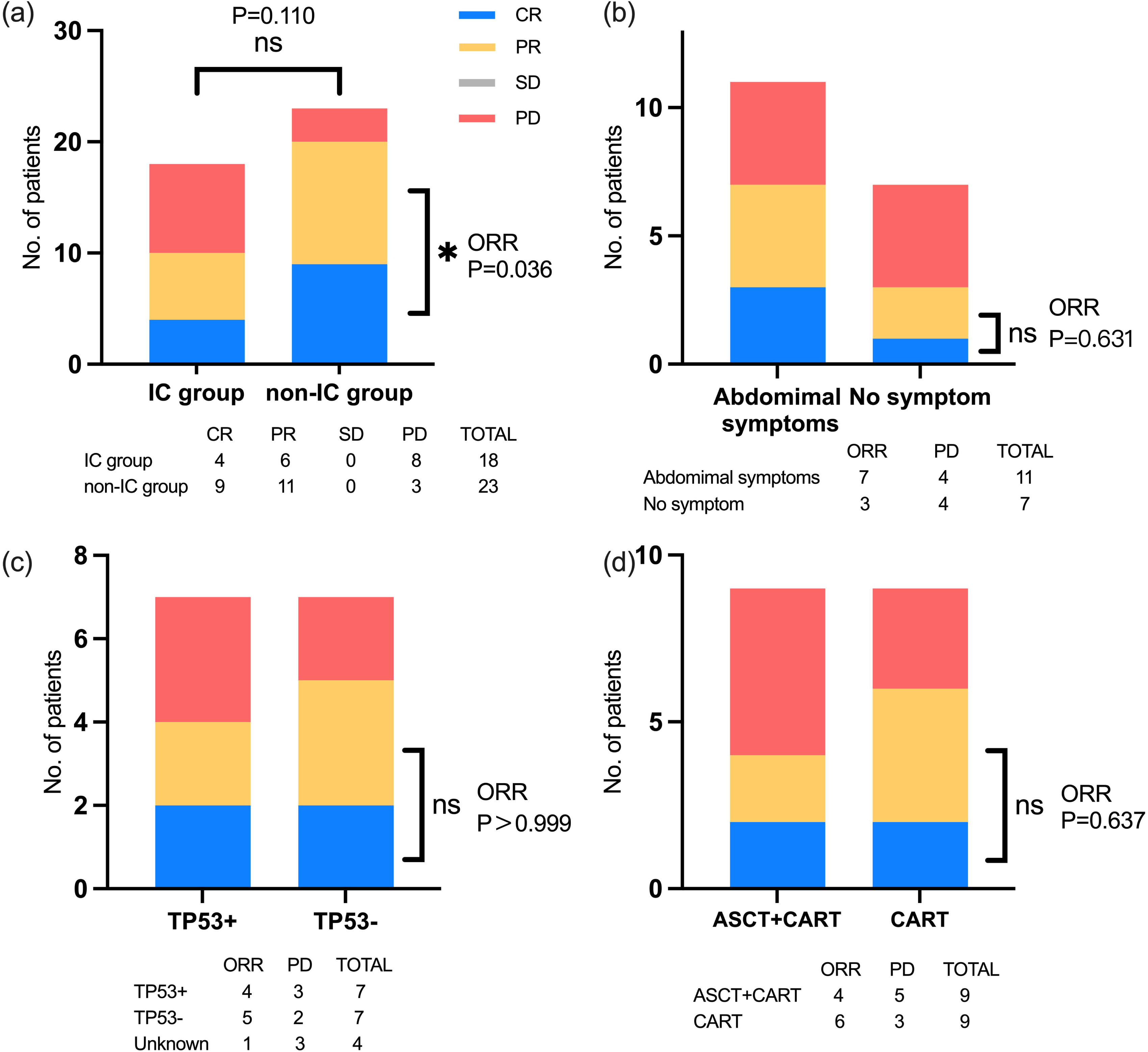
Three-month therapeutic responses. **(a)** IC versus non-IC groups: ORR 55.6% versus 87.0% (*P=*0.036). **(b)** Symptomatic versus asymptomatic subgroups: ORR 63.6% (7/11) versus 42.9% (3/7; *P=*0.110). **(c)** TP53-mutant (TP53+) versus TP53-wild-type (TP53-): ORR 57.1% versus 71.4% (P > 0.999). **(d)** ASCT+CAR-T versus CAR-T alone: ORR 44.4% (4/9) versus 66.7% (6/9; *P=*0.637). CR: Complete remission; PR: Partial remission; SD: Stable disease; PD: Progressive disease; ORR: Objective response rate; ASCT: Autologous hematopoietic stem cell transplantation; CAR-T: Chimeric antigen receptor T-cell therapy. *: P < 0.05; ns: Not significant.

### Adverse Reactions

Analysis of CAR-T-related toxicities revealed that grade 1 cytokine release syndrome (CRS) was the most common adverse event in both ileocecal (14/21, 66.67%) and non-ileocecal cohorts (14/23, 60.87%) **(Fig. 4A)**, with no significant intergroup difference in CRS severity distribution (*P=*0.565). Three ileocecal patients (14.29%) developed grade 3 CRS. One patient (4.76%) with diffuse large B-cell lymphoma (GCB subtype) and bone marrow involvement experienced grade 4 CRS accompanied by disseminated intravascular coagulation (DIC), grade 4 immune effector cell-associated neurotoxicity syndrome (ICANS), and septic shock (polymicrobial bacterial, fungal, and viral infections), ultimately succumbing to septic shock 20 days post-CAR-T infusion.

**Fig. 4.**
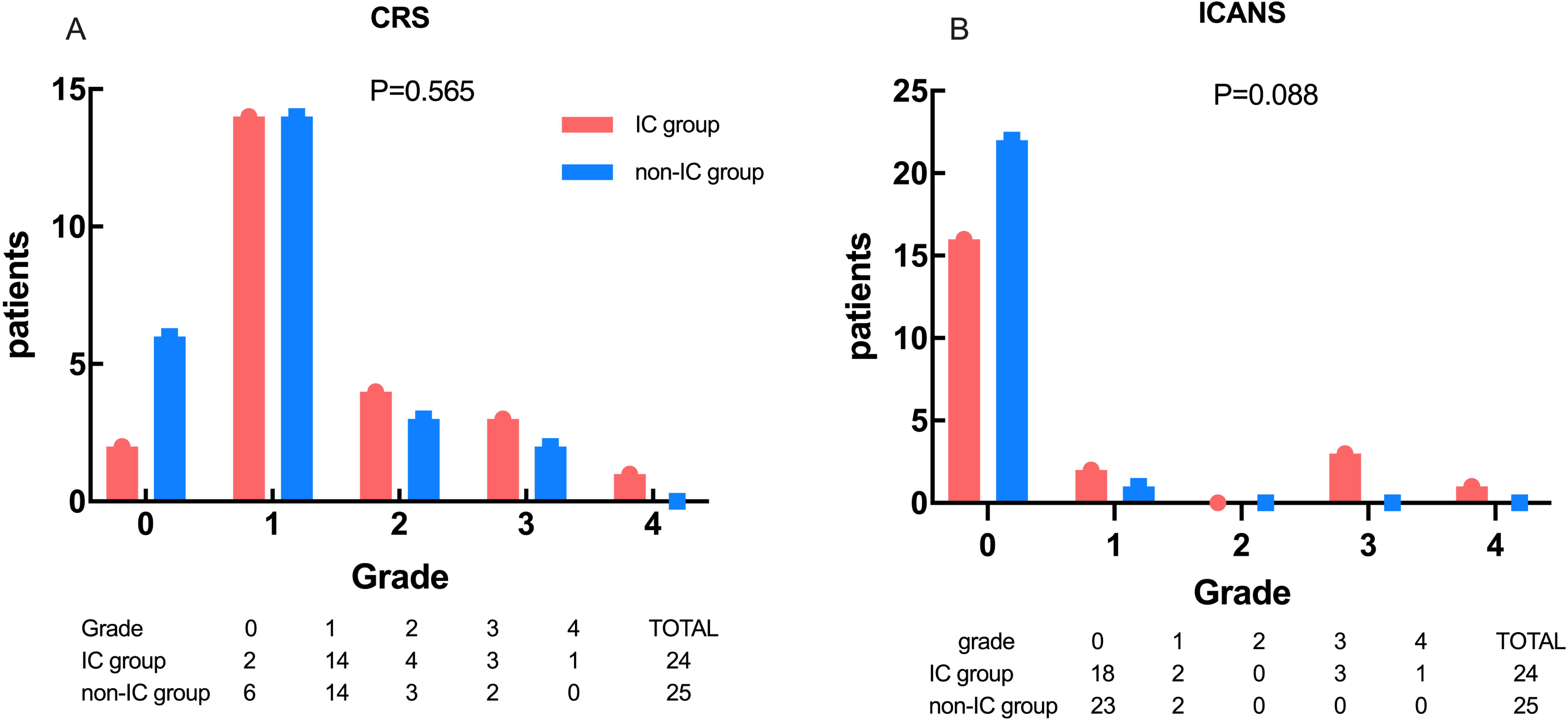
Adverse events after CAR-T therapy. **(A)** CRS severity distribution in ileocecal (IC) versus non-ileocecal (non-IC) groups (no significant difference, *P=*0.565). **(B)** ICANS severity distribution between groups (no significant difference, *P=*0.088). The x-axis represents toxicity grades, and the y-axis indicates the number of affected patients, with red and blue bars denoting IC and non-IC groups, respectively. In the IC cohort, 24 toxicity events were recorded: one patient (P18) received ASCT followed by CAR-T retreatment after relapse; one (P16) underwent ASCT+CAR-T post-progression; and one (P19) received two CAR-T courses followed by ASCT+CAR-T. The non-IC cohort reported 25 toxicity events, including two patients who received secondary CAR-T therapy after ASCT+CAR-T failure. IC: ileocecal region; non-IC: non-ileocecal region; CRS: cytokine release syndrome; ICANS: immune effector cell-associated neurotoxicity syndrome.

Immune effector cell-associated neurotoxicity syndrome (ICANS) was absent in 16 ileocecal (76.19%) and 21 non-ileocecal patients (91.30%), with no significant difference in severity between groups (*P=*0.088) **(Fig. 4B)**. Grade 3 ICANS occurred in three ileocecal patients (14.29%): one case of DLBCL (GCB subtype) with central nervous system involvement exhibited concurrent grade 3 CRS, demonstrated progressive disease (PD) at the 3-month assessment, and died from disease progression 5 months post-treatment; one Burkitt lymphoma patient showed PD at 3 months and discontinued therapy due to progression at 5 months; and one patient (4.76%) developed grade 4 ICANS. All four ileocecal patients with severe ICANS (≥grade 3) had concurrent CRS ≥grade 2. Non-ileocecal patients experienced only grade ≤1 ICANS.

### Survival Analysis

Survival outcomes are summarized in **Table 2**. As of the final follow-up date (March 7, 2025), the median follow-up duration was 33 months. Five patients died during the study period, including four due to disease progression and one from sepsis. Two patients were lost to follow-up after treatment discontinuation due to disease progression. Seven patients with <1 year of follow-up were excluded from long-term survival analyses.

**Table 2.** Survival outcomes in IC and non-IC groups after CAR-T therapy.

|  | Median survival ( m ) |  | Log-rank | P value |
| --- | --- | --- | --- | --- |
|  | IC Group<br>(n=21) | non-IC Group<br>(n=23) | HR ( 95%CI ) |  |
| 1year-OS | undefined | 64 | 1.303(0.4470-3.795) | 0.613 |
| 3year-OS | 10.5 | 27 | 1.145(0.4518-2.901) | 0.766 |
| 1year-PFS | 3.5 | undefined | 2.593(1.094-6.146) | 0.012* |
| 3year-PFS | 3.5 | 27 | 3.179(1.412-7.156) | 0.0003*** |

The 1-year overall survival (OS) rate was 50.0% (95% CI: 23.8-76.2), with a median 3-year OS of 10.5 months and a cumulative incidence of 50% (95% CI: 23.8-76.2) **(Fig. 5A)**. The non-ileocecal group exhibited a median 3-year OS of 27 months, with no significant differences in 1-year or 3-year OS between groups (*P=*0.697; *P=*0.766). The median progression-free survival (PFS) for the ileocecal cohort was 3.5 months, with 1-year and 3-year PFS rates of 18.8% (95% CI: 4.7-40.2) and 0%, respectively **(Fig. 5B)**. In contrast, the non-ileocecal group demonstrated a significantly superior median 3-year PFS of 27 months, with both 1-year and 3-year PFS rates markedly higher than those of the ileocecal group (*P=*0.012 and *P=*0.0003).

**Figure. 5.**
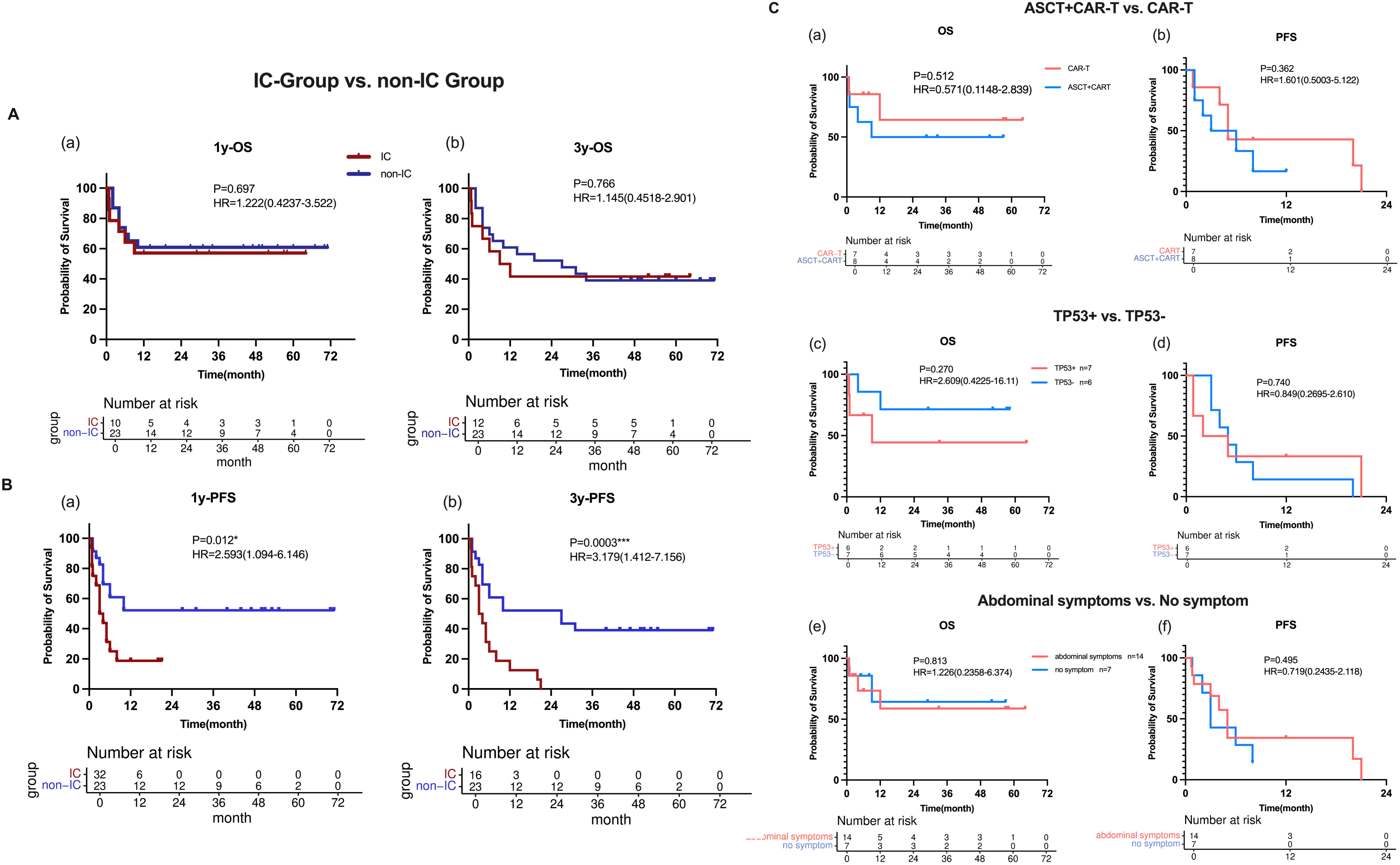
Survival outcomes. **A:** OS comparison between ileocecal (IC) and non-ileocecal (non-IC) groups: **(a)** 1-year OS; **(b)** 3-year OS. **B:** PFS comparison: **(a)** 1-year PFS; **(b)** 3-year PFS. **C:** Subgroup analyses: **(a)** OS for ASCT+CAR-T versus CAR-T alone; **(b)** PFS for ASCT+CAR-T versus CAR-T alone; **(c)** OS for TP53+ versus TP53−; **(d)** PFS for TP53+ versus TP53−; **(e)** OS for symptomatic versus asymptomatic subgroups; **(f)** PFS for symptomatic versus asymptomatic subgroups. The x-axis represents follow-up time (months), and the y-axis indicates survival probability. Downward curve inflections denote event occurrences (death or progression). Vertical tick marks indicate censored data (loss to follow-up or event-free at study termination). OS: overall survival; PFS: progression-free survival; IC: ileocecal region; non-IC: non-ileocecal region; ASCT: autologous hematopoietic stem cell transplantation; CAR-T: chimeric antigen receptor T-cell therapy. Status: 1=event occurred; 0=alive.

Subgroup analyses revealed no significant differences in OS or PFS based on abdominal symptom presence, TP53 mutation status, or prior autologous stem cell transplantation (ASCT) before CAR-T therapy **(Fig. 5C)**.

### Kinetics of CAR-T Cells

CAR-T cell kinetics were quantified via digital PCR. Ileocecal patients exhibited significantly higher CD19 peak copy numbers (mean: 67,290; range: 69-288,940; *P=*0.009) and CD19 AUC_0-last_ (mean: 451,592; range: 384-1,628,799; *P=*0.019) compared to non-ileocecal patients**(Fig. 6A)**. The mean time to CD19 peak was 8.2 days (range: 0-13), with a mean persistence duration of 49.43 days (range: 0-253).

**Figure. 6.**
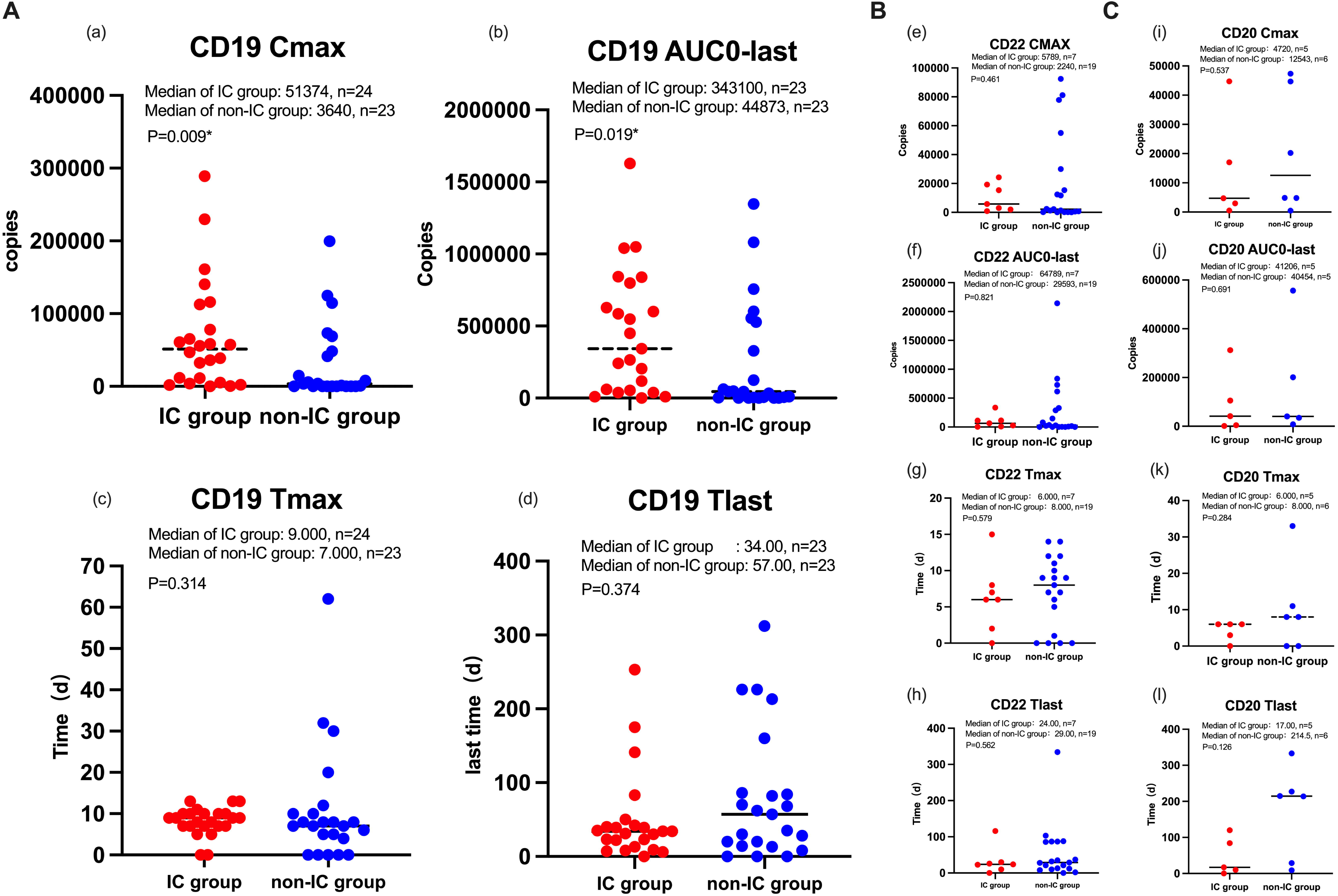
Kinetics of CAR-T Cells. **A.** CD19-targeted kinetics: **(a)** CD19 Cmax; **(b)** CD19 AUC0-last; **(c)** CD19 Tmax (time to peak copy number); **(d)** CD19 Tlast (duration of detectable copies). **B.** CD22-targeted kinetics: **(e)** CD22 Cmax; **(f)** CD22 AUC0-last; **(g)** CD22 Tmax; **(h)** CD22 Tlast. **C.** CD20-targeted kinetics: **(i)** CD20 Cmax; **(j)** CD20 AUC0-last; **(k)** CD20 Tmax; (l) CD20 Tlast. Data are presented as median with interquartile range. Intergroup comparisons were performed using Mann-Whitney U tests, with significance denoted as P < 0.05, *P < 0.01, **P < 0.001. AUC: Area Under the Curve.

For CD22-targeted CAR-T cells, ileocecal patients showed comparable peak copy numbers (mean: 10,087; range: 906-24,266; *P=*0.461) and AUC_0-last_ (mean: 94,339; range: 2,895-335,266; *P=*0.821) to non-ileocecal patients **(Fig. 6B)**. The mean time to CD22 peak (6.29 days; range: 0-15) and persistence duration (32.71 days; range: 0-116) also showed no intergroup differences (*P=*0.579 and *P=*0.562, respectively).

In five patients receiving ASCT followed by CAR-T therapy, CD20-targeted CAR-T cells demonstrated a mean peak copy number of 13,997 (range: 526-44,737) and AUC_0-last_ of 92,791 (range: 1,490-311,940) **(Fig.6C)**. The mean time to CD20 peak (4.2 days; range: 0-6) and persistence duration (46.2 days; range: 0-120) were comparable to the non-ileocecal group (*P=*0.537,*P=*0.591, *P=*0.284, and *P=*0.126 for peak, AUC, time to peak, and duration)

### Multivariate Analysis

Univariate Cox regression analysis of overall survival (OS) and progression-free survival (PFS) in ileocecal patients identified potential prognostic factors (P < 0.1), which were subsequently included in multivariate analysis (**Figs. 7,8**). Factors with P < 0.05 in multivariate Cox regression were considered independent predictors. For PFS, multivariate analysis revealed two independent predictors: 3-month post-CAR-T treatment response and pretreatment lines (**Table 3**). Due to multicollinearity (variance inflation factor, VIF > 10 for all variables) and limited sample size, Lasso regression was employed for OS multivariate analysis (**Table 4**). Regression coefficients > 0 indicated independent prognostic significance. Lasso regression identified four independent predictors of OS: 3-month post-CAR-T treatment response, time to B-cell recovery, baseline absolute lymphocyte count, and cumulative PD-1 inhibitor usage (**Fig. 9**).

**Figure. 7.**
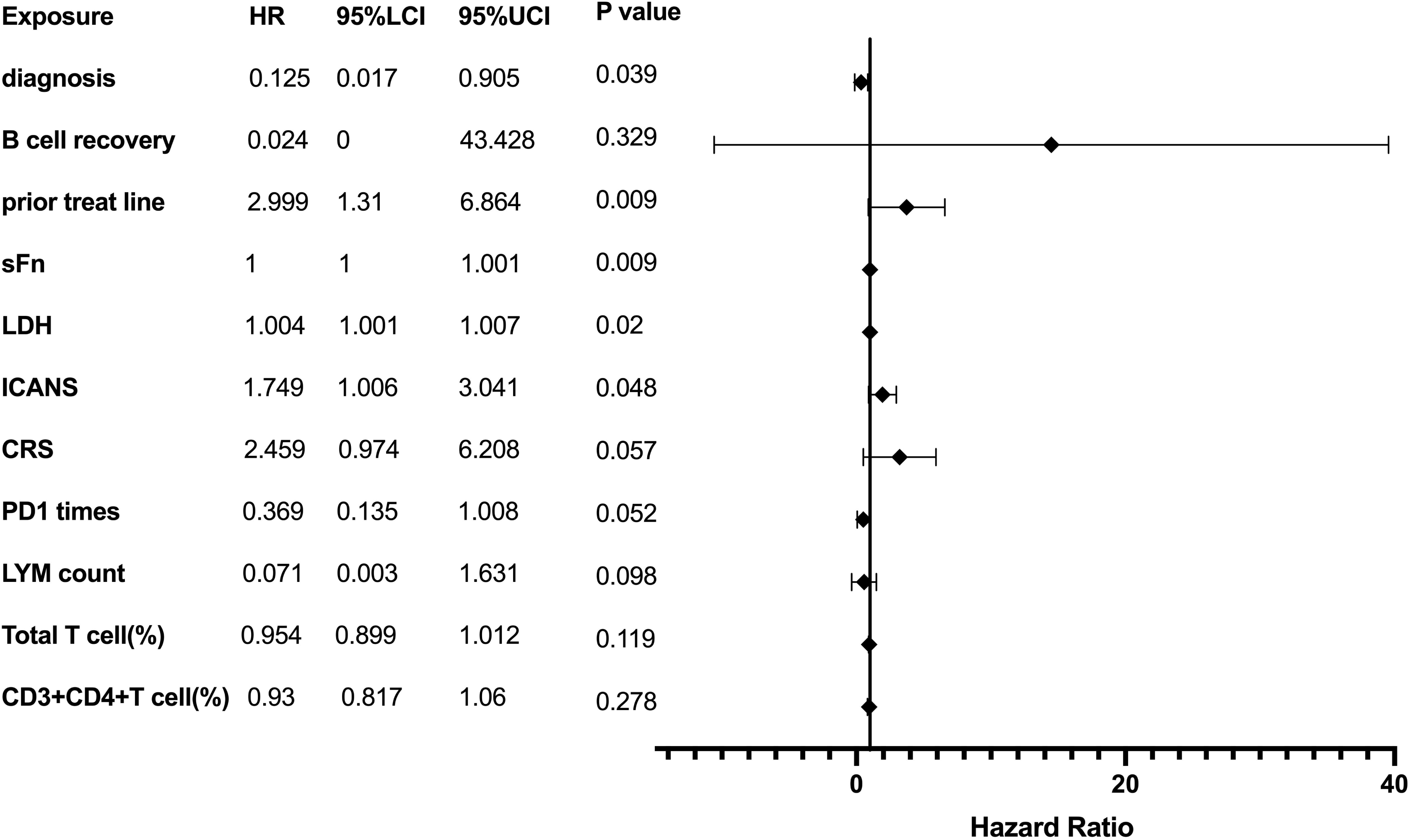
Univariate analysis of OS. The x-axis represents hazard ratios (HR) with a scale of 0-40; the vertical dashed line indicates HR=1 (no effect). Black dots denote HR values, horizontal lines represent 95% confidence intervals (CI). Significance was defined as CI excluding 1 with *P* < 0.05. LYM count: Baseline absolute lymphocyte count pre-CAR-T; PD1 times: Cumulative PD-1 inhibitor administrations post-CAR-T; sFn: Serum ferritin; Total T cell (%): Proportion of total T cells in lymphocyte subsets at 1 month post-CAR-T; CD3+CD4+ T cell (%): Proportion of CD3+CD4+ T cells at 1 month post-CAR-T.

**Figure. 8.**
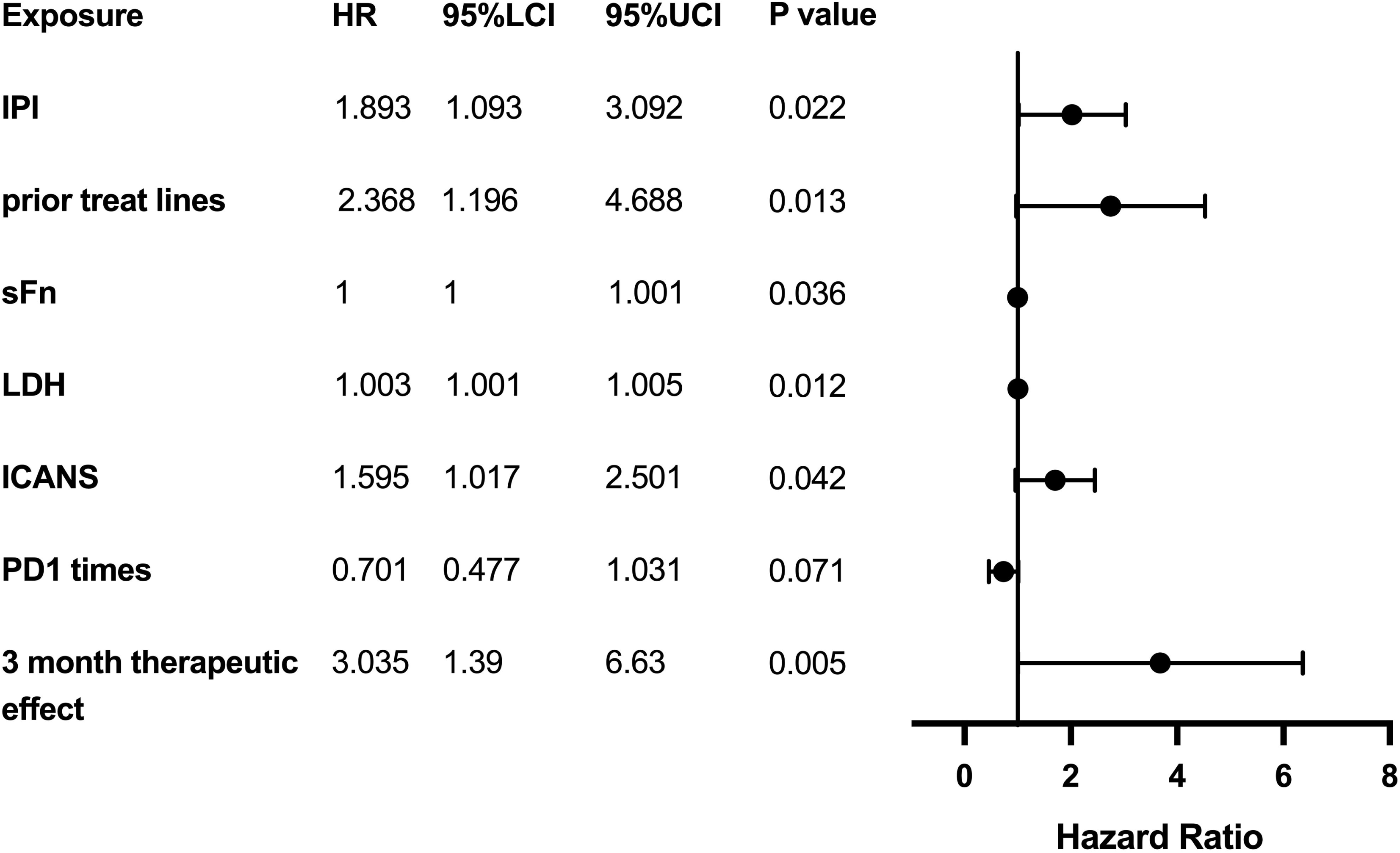
Univariate analysis of PFS. Formatting and significance criteria mirror Figure 8. 95% LCI: 95% lower confidence interval; 95% UCI: 95% upper confidence interval.

**Figure. 9.**
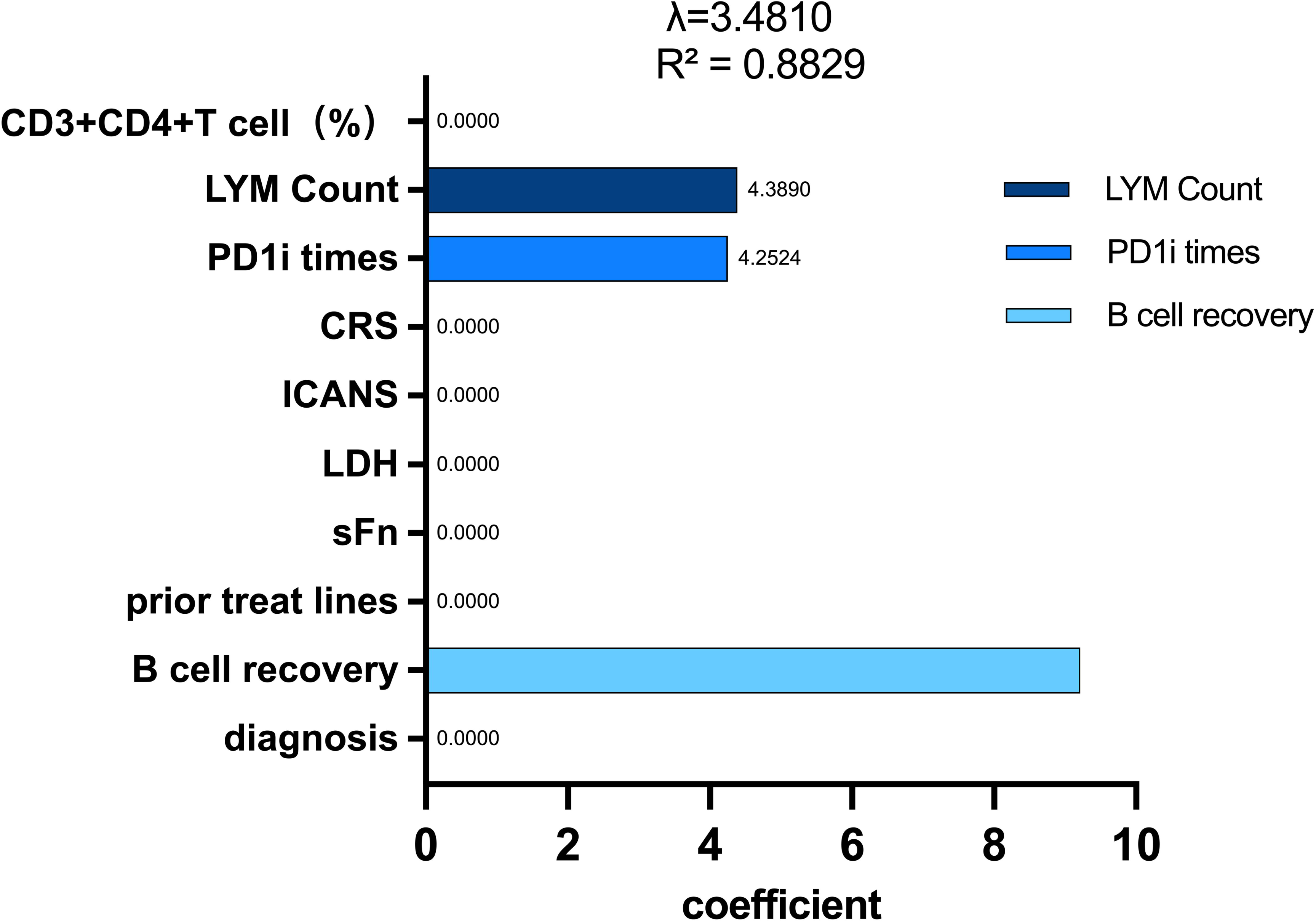
Lasso regression multivariate analysis of OS. The x-axis lists variable names; the y-axis displays regression coefficients. Positive coefficients indicate protective factors (all variables in this analysis). Dots represent coefficient magnitudes.

**Table 3.**
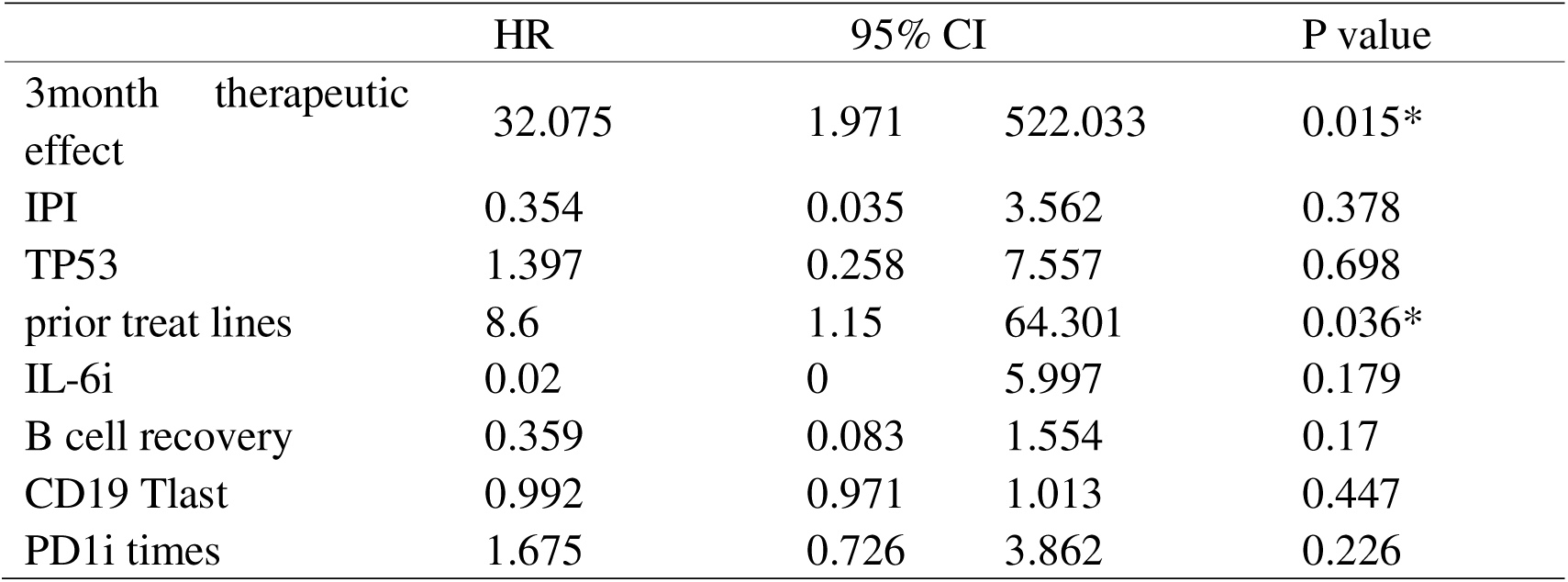
COX regression Multi-factor analysis of PFS.

|  | HR | 95% CI |  | P value |
| --- | --- | --- | --- | --- |
| 3month therapeutic effect | 32.075 | 1.971 | 522.033 | 0.015* |
| IPI | 0.354 | 0.035 | 3.562 | 0.378 |
| TP53 | 1.397 | 0.258 | 7.557 | 0.698 |
| prior treat lines | 8.6 | 1.15 | 64.301 | 0.036* |
| IL-6i | 0.02 | 0 | 5.997 | 0.179 |
| B cell recovery | 0.359 | 0.083 | 1.554 | 0.17 |
| CD19 Tlast | 0.992 | 0.971 | 1.013 | 0.447 |
| PD1i times | 1.675 | 0.726 | 3.862 | 0.226 |

**Table 4.** Multi-col linearity of OS.

| Exposure | Significance | Tolerance | VIF |
| --- | --- | --- | --- |
| 3m therapeutic effect | -0.326 | 0.066 | 15.093 |
| PD1i times | 0.651 | 0.008 | 132.28 |
| CD19 Tlast | 0.043 | 0.015 | 65.175 |
| lymphocyte Count<br>( first day of ASCT/CAR-T ) | -0.892 | 0.019 | 51.659 |
| diagnosis | 0.254 | 0.203 | 4.933 |
| sFn | -0.787 | 0.02 | 50.787 |
| LDH | 1.394 | 0.014 | 74.026 |
| ICANS | -0.655 | 0.024 | 41.794 |
| CRS | 1.315 | 0.017 | 59.473 |
| CD3+CD4+T cell ( % ) | -1.114 | 0.007 | 140.696 |
| total T cell ( % ) | 2.439 | 0.005 | 220.145 |
| B cell recovery | -1.621 | 0.011 | 93.922 |
| prior treatment line | 0.575 | 0.011 | 90.743 |

Abbreviation: VIF, Variance inflation factor

## DISCUSSION

Chimeric antigen receptor T-cell (CAR-T) therapy has demonstrated groundbreaking efficacy in relapsed/refractory B-cell lymphomas^[11-13]^. The 5-year follow-up data from the ZUMA trial revealed that Axicabtagene Ciloleucel achieved a complete response (CR) rate of 58%, with a median overall survival (OS) of 25.8 months and an estimated 5-year OS rate of 42.6%^[14]^. However, subgroup analyses focusing on intestinal lymphomas remain scarce^[15]^. This study evaluates the efficacy and safety of CAR-T therapy in 21 patients with relapsed/refractory ileocecal lymphoma, providing critical insights into therapeutic mechanisms and clinical decision-making.

The ileocecal region represents the second most common site of primary gastrointestinal lymphoma (20-30%)^[1,2,8]^ and the most frequently involved site for primary intestinal lymphoma (20.3-39.8%)^[1-4]^, predominantly comprising diffuse large B-cell lymphoma (DLBCL) and mucosa-associated lymphoid tissue (MALT) lymphoma^[2]^, with rare T-cell or NK-cell subtypes^[9]^. Consistent with literature, 85.7% of our cohort had DLBCL, and survival analysis revealed that Burkitt lymphoma exhibited poorer outcomes compared to DLBCL. At initial diagnosis, 80.95% of patients presented with stage IV disease, often involving bone marrow (14.29%) or central nervous system (CNS) involvement (4.76%), while 71.43% exhibited B symptoms, underscoring the aggressive clinical behavior of ileocecal lymphoma. The mean of 2.3 prior treatment lines before CAR-T therapy highlights the propensity for chemoresistance following standard therapies. Early retrospective studies on ileocecal lymphoma similarly reported that conventional therapies such as surgery, radiotherapy, or chemotherapy failed to improve long-term survival^[16]^.

Next-generation sequencing analysis of 15 ileocecal lymphoma patients revealed a gene mutation profile; TP53 mutations occurred at a rate of 42.86%, representing the most frequent mutation type in this cohort, aligning with the previously reported trend of high TP53 mutation frequency in ileocecal lymphoma^[17]^. Notably, the rate of CD79B mutations/aberrant expression in this cohort was 21.4% (3/14), significantly higher than the reported low expression or absence rate of CD79B (∼8%) in primary gastrointestinal lymphomas^[18]^; this difference suggests mutational heterogeneity between ileocecal lymphoma and lymphomas originating from other gastrointestinal sites or extranodal locations.

Furthermore, TP53 mutations in this study frequently co-occurred with mutations in EP300, KMT2D, and TET2; inactivation of these genes impairs histone acetylation, histone methylation, and DNA methylation, respectively, leading to epigenetic dysregulation and subsequent blockade of B-cell differentiation^[19-21]^. Concurrently, CD79B mutations constitutively activate the B-cell receptor (BCR) signaling pathway and its downstream NF-κ B pathway^[18]^. This indicates that ileocecal lymphomagenesis involves the cooperative dysregulation of multiple pathways, including TP53 inactivation, epigenetic dysregulation, and aberrant BCR signaling activation.

Although TP53 and KMT2D mutations are established contributors to chemotherapy resistance and poor prognosis in lymphoma^[19, 22-24]^-potentially explaining the poor initial response to standard therapy and frequent progression to relapsed/refractory disease observed in TP53-positive patients in this cohort-subsequent subgroup analysis demonstrated no significant impact of TP53 mutation status on the 3-month objective response rate (ORR), progression-free survival (PFS), or overall survival (OS). A similar observation was reported by Edit Porpaczy et al. ^[25]^. Jing Pan et al. found that TP53 mutations or deletions in lymphoma patients can lead to CD19-negative antigen escape, increasing relapse rates after CD19-directed CAR-T therapy^[26]^; in this study, the application of sequential "cocktail" therapy using dual-target (CD19/22) and triple-target (CD19/20/22) CAR-T cells substantially reduced the likelihood of tumor immune evasion through single antigen (CD19) loss, which has been shown to significantly reduce relapse rates post-CAR-T therapy^[27]^.

This study detected that ileocecal lymphoma patients exhibited significantly higher peak expansion levels and AUC0-last of peripheral blood CD19 CAR-T cells compared to the control group; however, the objective response rate (ORR) at 3 months post-CAR-T therapy (55.56%) was significantly lower than that in the control group (86.96%), suggesting that while CAR-T cell expansion efficiency was superior in ileocecal patients, the robust expansion did not translate into durable remission, potentially due to immune factors inhibiting CAR-T cell function or accelerating CAR-T cell exhaustion in vivo.

First, alterations in gut microbiota and the immune microenvironment are significant factors in lymphomagenesis and impact CAR-T efficacy and toxicity. Release of the gut microbiota metabolite lipopolysaccharide (LPS) can interact with tumor necrosis factor (TNF), enhancing the NF-κB pathway via TLR4 signaling, leading to malignant proliferation of intestinal B cells and subsequent lymphoma development; short-chain fatty acids (SCFAs, e.g., butyrate) promote regulatory T cell (Treg) function and inhibit lymphomagenesis, while ursolic acid A demonstrated immunomodulatory and anti-proliferative effects on lymphoma cells in vitro^[28]^. Uribe-Herranz M et al.^[29]^ found that altered gut microbiota in acute B-lymphoblastic leukemia patients influenced CAR-T cell expansion efficiency, and oral vancomycin administration modulated the gut microbiota, subsequently regulating T cell immunity and enhancing CAR-T cell expansion and treatment outcomes. Another study^[30]^ similarly reported that higher abundance of specific gut microbes (e.g., Bifidobacterium, Clostridium) correlated with milder cytokine release syndrome and improved treatment response during CAR-T therapy. Concurrently, studies found that modulating cytokine expression within the microenvironment and targeting immunosuppressive cells enhanced CAR-T cell function and improved patient outcomes^[31].^ This study only assessed CAR-T cell expansion in peripheral blood; further exploration is warranted regarding the correlation between CAR-T cell expansion efficiency within the ileocecal tissue and the gut microbiota/microenvironment.

Second, gut-associated lymphoid tissue (GALT) constitutes the core region for intestinal immune responses; the ileocecal GALT lamina propria is rich in Peyer’s patches (PP)^[32]^. Unlike the colorectal lamina propria, which is dominated by plasma cells, PP exhibit concentrated distributions of B cells and T cells. Early research found that PP efficiently adsorb the aromatic hydrocarbon carcinogen 3-methylcholanthrene(3-MC), playing a key role in inducing B-cell lymphomagenesis^[33]^. The mucosal epithelium of the ileocecal region harbors a significantly higher density of M cells compared to other gastrointestinal areas; these M cells mediate the active transport of external antigens from the intestinal lumen to dendritic cells within Peyer’s Patches, playing a critical role in mucosal immune surveillance^[34,35]^. M cells secrete IgA antibodies to neutralize pathogens and are also crucial for maintaining gut microbiota homeostasis^[36]^. However, the role of alterations in this structure in the toxicity and efficacy of CAR-T therapy for ileocecal lymphoma remains limited and warrants further investigation.

Current researchs on CAR-T therapy for ileocecal lymphoma are limited, with only a small number of studies available on gastrointestinal or intestinal lymphomas. A study including 14 patients with primary gastrointestinal DLBCLreported 6-month overall survival (OS) and progression-free survival (PFS) rates of 58.33% and 54.55%, respectively, following CAR-T therapy^[37]^. A previous multivariate analysis of 81 intestinal lymphoma patients found that ileocecal involvement was an independent prognostic factor associated with improved OS and event-free survival (EFS), while univariate analysis showed that the absence of perforation correlated with better OS ^[38]^. In this study, all 14 patients presented with gastrointestinal symptoms, primarily abdominal distension, intestinal obstruction, intestinal perforation, and bleeding; 4 patients required emergency surgery, suggesting that local tissue structural destruction may exacerbate tumor aggressiveness. However, the median PFS in the ileocecal group in this study was only 3.5 months, significantly lower than the 27 months in the non-ileocecal group. Both 1-year and 3-year PFS rates were significantly lower in ileocecal patients compared to non-ileocecal patients, but no difference in OS was observed; subgroup analysis revealed no significant difference in PFS or OS based on the presence or absence of abdominal symptoms. Concurrently, we did not find that surgical intervention improved patient survival, which deviates from the findings described in the aforementioned studies; this discrepancy is potentially attributable to the fact that patients in this study were all relapsed/refractory with high tumor burden, and differences in intestinal immune status may have led to CAR-T cell therapy resistance or reduced persistence, resulting in rapid disease progression. Second, the aforementioned studies primarily employed conventional treatments such as surgery, radiotherapy, and chemotherapy, whereas our approach was based mainly on CAR-T and autologous stem cell transplantation (ASCT), with different treatment modalities leading to divergent outcomes. Third, the small sample size in this study may have introduced statistical bias. The lack of difference in 1-year and 3-year OS suggests that although disease progression occurred shortly after CAR-T therapy, patients still achieved expected survival through various salvage therapies, including intensive chemotherapy, PD-1 inhibitors, second CAR-T infusions, or autologous hematopoietic stem cell transplantation.

In the analysis of CAR-T cell therapy-related toxicities, this study found that immune effector cell-associated neurotoxicity syndrome (ICANS) reactions were most frequently grade 0 and cytokine release syndrome (CRS) reactions were most frequently grade 1 in ileocecal lymphoma patients undergoing CAR-T therapy; no statistically significant differences were observed in the incidence of any grade of toxicity reaction between ileocecal and non-ileocecal patients, demonstrating favorable safety of CAR-T therapy in ileocecal lymphoma patients. One ileocecal patient with germinal center B-cell-like (GCB) subtype diffuse large B-cell lymphoma (DLBCL) involving bone marrow and central nervous system died due to grade 4 CRS, grade 4 ICANS, sepsis, and multi-organ failure, demonstrating that disease progression pre-transplantation and high tumor burden in the patient may exacerbate CAR-T toxicity^[39]^.

In multivariate analysis, we identified suboptimal response at 3 months post-CAR-T therapy as an independent prognostic factor for both progression-free survival (PFS) and overall survival (OS), suggesting that precise clinical assessment at this timepoint is warranted to guide prognosis and inform subsequent consolidation or maintenance therapy selection. Concurrently, we found that a higher number of lines of prior therapy was an independent prognostic factor for inferior PFS, while lower absolute lymphocyte count pre-CAR-T, delayed B-cell recovery post-CAR-T, and fewer administrations of PD-1 inhibitors were independent prognostic factors for inferior OS. Stock S et al.^[31]^similarly reported concordant findings, demonstrating that detectable B cells and T-helper (Th) cells >200 μL were associated with superior OS and PFS, and reduced risks of infection and non-relapse mortality. Cherkassky et al.^[40]^found that PD-1 inhibitors significantly downregulated PD-1 expression within the microenvironment, thereby enhancing CAR-T cell function and improving patient survival. It should be noted that the 95% confidence intervals (CIs) for the aforementioned exposure factors were wide, suggesting sparse data or underlying heterogeneity; further validation in larger cohorts is required.

In conclusion, our results indicate that CAR-T therapy is a feasible and manageable salvage option for relapsed/refractory ileocecal lymphoma patients failing ≥2 prior lines. Although ileocecal patients exhibited significantly inferior objective response rates (ORR) and PFS compared to non-ileocecal counterparts, comparable OS underscores the potential of post-progression salvage therapies (e.g., chemotherapy, PD-1 inhibitors, or secondary CAR-T/ASCT) to mitigate early treatment failure. Early response assessment (3-month evaluation) and B-cell recovery kinetics should serve as pivotal biomarkers for personalized management. Optimizing CAR-T timing (e.g., frontline application) may enhance long-term outcomes in this high-risk population. These insights provide a theoretical foundation for incorporating CAR-T into the salvage paradigm for ileocecal lymphoma. The clinical recommendation for patients with ileocecal lymphoma following CAR-T treatment, as indicated in this study, is to perform an early assessment at the three-month mark to predict PFS and OS. Additionally, the administration of immunomodulatory agents(such as PD-1 inhibitors), should be considered for maintenance therapy to enhance long-term patient survival outcomes. However, the single-center design, small sample size, and potential selection bias inherent to this study warrant validation through multicenter trials with expanded cohorts.

## Data Availability Statement

The data supporting the findings of this study are available from the corresponding authors upon reasonable request and subject to institutional and ethical guidelines.

## Authors’ Contributions

XZ, ZL and WW designed and directed the study. ZS, XC, FM, YC and ZZ collected the clinical data and conducted the follow-up. ZS contributed in data analysis and wrote the paper. All authors had full access to the primary study data and reviewed the manuscript.

## Acknowledgement

We sincerely thank the 21 patients and their families for participating in this study. We acknowledge the clinical teams at Tongji Hospital and Guangxi Medical University for patient care and data collection. Special gratitude to Wuhan Bio-Raid Biotechnology Co., Ltd. for providing investigational CD19/CD22 CAR-T cells. Technical support from laboratory staff for molecular analyses (NGS, dPCR) is appreciated. This work was supported by the National Natural Science Foundation of China (82270173) and Hubei Provincial Medical Youth Top Talent Project. We deeply thank Professors Xiaojian Zhu, Wang Wei, and Zhenfang Liu for their guidance.

## Patient and Public Involvement

Due to the retrospective nature of this study, which analyzed existing data from patients who had already received CAR-T cell therapy, patients were not directly involved in the setting of the research question, study design, or analysis. However, the outcomes of this research were chosen and prioritized based on our close clinical engagement with patients with relapsed/refractory B-cell malignancies, which highlighted overall survival, treatment-related toxicity, and predictors of long-term response as their greatest concerns. We are committed to disseminating the key findings of this study to the patient community. We will collaborate with our institutional patient support group and the Department of Hematology to produce a plain-language summary of the results. This summary will be made available through the hospital patient portal and shared with relevant patient advocacy organizations.

